# Exposing the need for a training programme in paediatric and adolescent gynaecology for paediatric and GP trainees

**DOI:** 10.1101/2020.05.26.20113449

**Authors:** EJ Cosgrave, JM Geraghty, AR Geoghegan

## Abstract

**Objective:** As paediatric and adolescent gynaecology (PAG) falls within the remit of paediatrics and gynaecology, training in both specialties is underdeveloped. There is a paucity of research investigating trainee knowledge of PAG, while postgraduate paediatric training demonstrates little focus in the field. Compounding this, a finite number of PAG specialists means clinical training is limited. We hypothesize that knowledge deficits exist among paediatric and GP trainees and that this has future implications for increased morbidity in girls.

**Design, Setting and Participants:** A structured questionnaire assessing PAG was distributed to forty paediatric and GP trainees in October 2019 in a tertiary paediatric hospital.

**Results:** 60% (24) incorrectly identified vulvovaginitis as candidal infection. 80% (32) were unable to identify labial adhesions. 62% (25) were unable to define menorrhagia. 100% (9) of GP trainees said they would prescribe the OCP compared with 51% (16) of paediatric trainees. 52% (21) did not consider STI screening when appropriate. 75% (30) believed genital warts invariably warrant referral to child sexual assault clinic. 70% (28) could not identify female genital mutilation. 60% (24) did not consider imperforate hymen as a cause of primary amenorrhoea. 67% (27) misdiagnosed lichen sclerosis.

**Conclusions:** A knowledge deficit among trainees was evident in relation to PAG conditions. Misdiagnosis and delayed treatment could lead to considerable increased morbidity for girls and we postulate that a key intervention which may prove effective in improving trainee competency in PAG lies in the introduction of a structured training curriculum for all clinicians involved in PAG practice.

## Introduction

Paediatric and adolescent gynaecology (PAG) can be anxiety-provoking for paediatricians, GPs and even some gynaecologists. Adult women’s health generally suffers from less investment leaving little space for research and education into conditions specific to prepubertal and teenage girls. For example, regarding studies on medications, animals researched are often exclusively male despite differences in drug metabolism in females.^1^ Women are less likely to be included in phase one clinical trials^2^ and medical issues specific to women and girls are less frequently studied. On a PubMed literature search in May 2020, the phrase ‘testicular torsion’ yielded 3,516 results versus 1,938 results for the term ‘ovarian torsion’.^3,4^ While there are a multitude of studies that *do* pertain to women and girls health very appropriately, some, in the authors’ opinion are not particularly useful.

Inspiration for this study arose from a paper which was published in a peer-reviewed journal in 2013 and subsequently gained significant attention on social media. This study reported on attractiveness levels of women with recto-vaginal endometriosis. ^5^ Furthermore, anecdotally, it was noticed that many prepubertal girls are diagnosed with candidiasis rather than vulvovaginitis and are subsequently treated incorrectly. Finally, in one instance a 14 year old girl was referred to a child sexual abuse clinic after she was told by a doctor examining her for itch, that she could not be a virgin based on the appearance of her hymen, despite the fact that she had reported no sexual intercourse/abuse. Her hymen appeared normal.

There is a paucity of research pertaining to trainee knowledge of paediatric and adolescent gynaecology. Published data from the UK does report on limitations in this area, however this research primarily focuses on obstetric trainee knowledge and mainly assesses knowledge of services available rather than exploring ability or confidence among obstetric trainees in relation to diagnosing and treating conditions. ^6^ The British Society of Paediatric and Adolescent Gynaecology (BritSPAG) have developed guidelines for clinicians managing relevant conditions but there is still no reference to paediatric gynaecology on either the Irish (RCPI) or UK (RCPCH) paediatric specialist training curricula. Once qualified, this lack of trainee PAG experience translates to uncertainty in the field and inappropriate onward referral to already limited specialist PAG services.

We hypothesize that paediatricians and general practitioners are not confident at diagnosing and managing common paediatric and adolescent gynaecology conditions or recognising serious ones and that this has implications for increased morbidity in girls.

## Methods

General practice (GP) and paediatric trainees were surveyed in a tertiary Irish paediatric hospital over a two-week period in October 2019 with a ten-question questionnaire to assess their knowledge of PAG. The survey consisted of questions relating to common presentations such as vulvovaginitis, labial adhesions, lichen sclerosis and menstrual dysfunction. It also assessed knowledge of rare but serious presentations such as Female Genital Mutilation (FGM), sexually transmitted infections (STIs) and haematometrocolpus. Answers were predefined from the authors’ local hospital guidelines and Royal Children’s Hospital Melbourne guidelines.^7^ The questionnaire included fictional case vignettes and diagrams and was colour printed on A4 paper. It was provided to candidates face-to-face by the researchers to allow for opportunistic recruitment of participants and to prevent possible cheating. Two pilot surveys were trialled in advance with a male and female participant. Trainees entered written answers on designated answer spaces provided on the survey. Trainees were told not to include any personal details such as their name on the survey to preserve anonymity when the researchers were analysing the data.

All three authors who are female were senior paediatric or GP trainees at the time of the study and approached prospective participants who were colleagues to seek their consent for involvement in the study. Trainees were approached in the workplace on the wards or in their teams’ office space. Fifty doctors were asked if they would like to participate. Ten candidates who were offered the chance to take part refused, all of whom cited an overburdened workload as the cause for refusal. Candidates were informed that any questions that they asked of the researcher would not elicit a response and no prompts were provided. While the survey was being completed, the researcher physically moved away from the candidate but stayed within their view. No time limit was placed on the candidate in which to complete the survey. Surveys were collected immediately following completion to ensure a true representation of baseline knowledge and placed in a folder without reviewing content. Participants were at this stage offered the correct answers if they wished and given the opportunity to provide feedback or ask questions. No repeat surveys were performed. Content was subsequently analysed by all three authors and data coded on an Excel spreadsheet. As most answers were binary i.e. either right or wrong, correct answers were coded as 0 while incorrect answers were coded as 1. One qualitative question was included in relation to trainees’ opinion regarding the prescription of the oral contraceptive pill and consistent themes were extracted and quoted in the results.

## Results

Forty trainees were surveyed. 70% (28) were female. 23% (9) were GP trainees and 77% (31) were paediatric trainees. 55% (22) were senior house officers and 45% (18) were registrars. A fictional case vignette was used to assess vulvovaginitis knowledge. 60% (24) of respondents incorrectly identified vulvovaginitis as a candidal infection and suggested antifungal treatment as the management plan. Of those who did identify vulvovaginitis correctly, 57% (14) listed the correct first line treatment plan of loose cotton underwear, avoiding soaps and bubble baths and using vinegar baths.

An image was used to assess trainees’ ability to identify labial adhesions. 80% (32) were unable to identify them and proposed treatment was thus incorrect. Of those who correctly identified labial adhesions 62% (5) were aware of the correct treatment i.e. Vaseline applied roughly, oestrogen creams or surgical management in some severe cases.

Menorrhagia is quite common in adolescents, but trainees had variable levels of knowledge and experience in treating it. 62% (25) were unable to correctly define menorrhagia and 55% (22) were unaware that it can take up to 6 years for cycles to regulate. Non-hormonal treatment options such as tranexamic acid and mefenamic acid were poorly recognised and although 98% (39) of respondents correctly identified the oral contraceptive pill (OCP) as a potential treatment only 63% (25) responded that they would feel comfortable prescribing it. 100% (9) of GP trainees said they would prescribe the oral contraceptive pill compared with 51% (16) of paediatric trainees. The main obstacles reported were ‘lack of gynaecology knowledge’, ‘lack of experience’ and ‘unfamiliarity with the oral contraceptive pill’s dosing and side effect profile’.

Rarer conditions were also included. 52% (21) did not consider sexually transmitted infection (STI) screening in the fictional case of a symptomatic child. However, 80% (32) of trainees did consider Herpes Simplex Virus as a cause of genital ulceration. 75% (30) of candidates suggested that genital warts invariably warrant referral to child sexual assault (CSA) clinic and were not aware of the concept of autoinoculation in younger children.

When candidates were shown an unlabelled diagram of types 1-3 of Female Genital Mutilation (FGM) 70% (28) could not identify it correctly.

Other findings included that 60% (24) did not consider imperforate hymen as a cause of primary amenorrhoea and 67% (27) misdiagnosed lichen sclerosis. Overall anatomical knowledge was good but polycystic ovarian syndrome was incorrectly suggested as a diagnosis by 20% (8).

## Discussion

Although initially, common presentations such as labial adhesions and menorrhagia may seem innocuous, misdiagnosis and delayed treatment can result in significant morbidity for girls and can frequently prompt inappropriate referrals to a limited PAG specialist service.

Unidentified labial adhesions can lead to recurrent urinary tract infections (UTIs) which in severe cases can lead to long-standing renal parenchymal disease.^8^ It is clear that trainees are not adept at identifying labial adhesions and although not directly assessed, we suspect that this may be due to the fact that standard practice among trainees likely does not include genital examination in the case of girls who present with recurrent UTIs. Non-obstructing labial adhesions often disappear during puberty once oestrogenisation of the vulva occurs. However, in the event of a labial adhesion disrupting effective urination by occluding the urethra, intervention with methods suggested above is necessary.

Menorrhagia is quite common in adolescents with 37% experiencing it. Paediatric trainees in particular had difficulty managing menorrhagia. In 2017, Torres et al reported on the impact on adolescents’ quality of life due to heavy menstrual bleeding; 50% of adolescents missed school, 80.4% missed physical education and 65.2% missed outdoor activities or parties. ^9^ Reluctance among clinicians to engage with girls in relation to the full spectrum of treatment options available to them is no doubt impacting on their quality of life and it is clear that further education is warranted. However, unless adolescent medicine departments are routinely established it is unlikely that paediatricians will take on this role as it requires significant training in contraception that is already delivered to GPs (albeit the training GPs receive generally pertains to prescribing contraception for adults). Whether or not paediatricians wish to develop the skills necessary to enable them to offer adolescents with menorrhagia the OCP, it is an important question to address collaboratively to ensure optimal health for this cohort.

Another point of interest that arose from this survey was the incorrect suggestion of polycystic ovary syndrome (PCOS) as a diagnosis, particularly in questions relating to menorrhagia and haematometrocolpus. Anecdotally, in the authors’ experience, many adolescents are referred or diagnosed with PCOS incorrectly when ovarian cysts are found on ultrasound incidentally. Although the presence of polycystic ovarian morphology is included as a key diagnostic criterion of PCOS in adults, it is currently not recommended for the diagnosis of adolescents. ^10^ In fact the actual criteria for diagnosis of PCOS in teenagers requires demonstration of ovulatory dysfunction and androgen excess i.e. oligomenorrhoea or amenorrhoea and hirsutism or persistent acne. Biochemical evidence of PCOS includes persistent elevation of testosterone levels and high luteinizing hormone (LH). Generally, the most effective treatment for PCOS is lifestyle modifications in obese adolescents but in some cases the OCP and metformin are prescribed to regulate menstrual cycles, improve acne and hirsutism and to treat insulin resistance.

The topics chosen for this study included common presentations but also rarer, more serious conditions. While we would not expect non-experts to know how to manage these more unusual diagnoses i.e. STIs or FGM, missing them can lead to life-altering consequences including infertility, chronic pain, dyspareunia, recurrent UTIs, issues with childbirth and even death. Female Genital Mutilation (FGM) is rare in Europe however with an increasingly diverse population it is an issue which doctors should have an awareness of and consider as it is an illegal practice in many countries. In an interagency statement published by the World Health Organisation (WHO) in 2008 FGM was described as a violation of human rights against girls and women, a form of gender discrimination, and a form of violence against girls^11^. The majority of FGM occurs in childhood, on average aged eight years. Outside the UK, 10% of girls die in the acute setting due to haemorrhage, shock and sepsis.^12^ The WHO estimates 100-140 million girls have been subjected to FGM worldwide and in a UK study by Forward in 2007 it was estimated that 21,000 girls were at risk of FGM in the UK.^13,14^ In addition to poor recognition of FGM via diagrammatic representation, overall trainee feedback was significant in that many felt unequipped to recognize and deal with a scenario in which a potential case presented.

Forensic gynaecology is beyond the scope of practice of most paediatricians and GPs and therefore knowledge in this area was not assessed, however due to the seriousness and social implications of the case referenced above which inspired the study, the authors feel it is necessary to comment on it. Genital injuries including hymenal tears are often absent following consensual/non-consensual intercourse in adolescents. In the event of an injury to the hymen the tissues frequently heal to leave no deficit or scar.^15^ It is usually impossible to comment on whether or not a girl is a virgin by looking at the hymen. Virginity testing still occurs around the world and has been shown to not only be inaccurate, but to cause physical, psychological and social harm to those examined.^16^

## Recommendations

Paediatric gynaecology does not appear on the Royal College of Paediatrics and Child Health (RCPCH) or Royal College of Physicians Ireland (RCPI) Paediatrics curriculum. The experience in Canada is comparable to Ireland, with trainees reporting limited paediatric gynaecology ability.^17,18^ We hypothesize that UK paediatric and GP trainees are also relatively unconfident in managing these conditions. The North American society for Paediatric and Adolescent Gynaecology (NASPAG) have created a Short and Long Curriculum to teach trainees core principles of this subspecialty area.^19^ The NASPAG recommend that the Short Curriculum is delivered during adolescent medicine rotations for paediatric residents which are an established part of training in the United States. Use of the Short Curriculum has been shown to be effective in improving self-reported basic knowledge in PAG among paediatric, GP and obstetric trainees.^20^ The programme provides a structured didactic curriculum covering basic topics including the ones assessed in this study that can be incorporated into a teaching programme. The Royal College of Paediatrics and Child Health (RCPCH) and Royal College of Physicians in Ireland (RCPI) Faculty of Paediatrics curricula are comprehensive and as a potential alternative to the Short Curriculum if it better suited UK and Irish training systems, we suggest the introduction of standardised paediatric and adolescent gynaecology study days for basic and higher specialist paediatric and GP trainees.

## Conclusion

In conclusion, this is the first Irish or UK study that confirms the suspected deficit in paediatric and GP trainee knowledge of paediatric and adolescent gynaecology (PAG) in keeping with very limited literature from other countries. We believe it is evident that trainee education pertaining to PAG needs to be expanded for paediatricians and GPs. As this subspecialty area falls within the remit of both paediatrics and gynaecology we postulate that trainees from gynaecology are also less confident in the area. Emphasis needs to be placed on both common presentations and rare but serious conditions, as the potential to miss more unusual gynaecological disorders was noted. To address this knowledge deficit locally, we plan to develop this into a quality improvement initiative by creating a gynaecology education programme in our paediatric hospital. We recommend and call for the inclusion of paediatric and adolescent gynaecology onto paediatric and GP training curricula.

## Data Availability

Survey data was collected on hard copies of questionnaires. They are in the care of AR Geoghegan at present. Participants were anonymous. Data was coded on an Excel spreadsheet with no personal participant information. This Excel spreadsheet is no longer available.

## Statement Page

**What is known about this topic:**

1. Paediatric gynaecology falls between two specialties - paediatrics and gynaecology, with many countries having limited access to specialist paediatric gynaecology.
2. Research from the UK and Ireland focuses on obstetric trainee’s lack of knowledge of paediatric gynaecology. There is very limited data on paediatrician or GP knowledge.
3. Paediatric gynaecology does not appear on the UK or Irish paediatric specialist training curriculum.

**What this study adds:**

- It confirms a suspected knowledge deficit among paediatric and gp trainees regarding paediatric and adolescent gynaecology, an area where a paucity of data exists.
- This study provokes thought regarding the unmet health needs of girls by discussing issues which are perhaps considered taboo, rare or innocuous.
- This study makes practical suggestions that could be developed and adapted further by the RCPCH and RCPI to increase trainee knowledge in this area.

## Acknowledgements

The authors wish to acknowledge Dr Geraldine Connolly who provided insight into the limitations of the current paediatric gynaecology service in Ireland.

